# Prevalence of anti-DENV IgG among routine Bangladeshi blood donors, and strategies for the future

**DOI:** 10.1101/2022.07.11.22277496

**Authors:** Ashraful Hoque, Sushanta Kumar Basak, A.B.M. Al-Mamun, Marufur Rahman, Kashfia Islam

**Author notes:** Corresponding author: Dr. Ashraful Hoque.

## Abstract

**Background:** Dengue is the most common arthropod-borne sickness worldwide, impacting at least 50 million people each year. The dengue virus has four primary serotypes. Infection with one serotype confers homotypic immunity but not heterologous immunity, and secondary infections may be more severe. Although blood transfusions and organ donations have also been observed, the Aedes aegypti mosquito is the primary vector for the transmission of dengue. Infection causes a continuum of clinical illness, from asymptomatic infection to dengue fever, DHF, and dengue shock syndrome (DSS).

**Aim:** To assess the presence of anti DENV IgG and anti DENV IgM antibodies specific to the four dengue serotypes in blood donor service donors and the importance of pre-donation screening in routine blood collection procedures.

**Method:** 3 mL of peripheral venous blood from 507 blood donors was collected in tubes with BD vacutainer gel tube for serum separation after epidemiological records were reviewed. After that, serum was separated and tests were performed by SD Bioline Dengue Duo. Participants in the study completed a social and epidemiological questionnaire that contained information such as age, gender, and dengue diagnosis.

**Result:** Out of the 507 blood samples that were taken, 473 (93.3%) came from male blood donors, while the remaining 34 (6.7%) belonged to female blood donors. The ratio of males to females is 13.91 to 1. The age range is 18–60 years, and the mean and standard deviation are both 27.7 and 6.5. 183 of the 507 samples produced anti DENV IgG positivity, while 324 did not. The ratio of positive to negative was 1.25:2.

**Conclusion:** According to the findings of this study, quantitative methods for determining the presence of anti-dengue antibodies or detecting the dengue virus in blood donors in endemic areas should be devised in order to ensure the quality of blood transfusions.

## Introduction

Dengue is a virus that is spread by mosquitoes and has quickly spread to all WHO regions in the past few years. Dengue virus is mostly spread by female Aedes aegypti mosquitoes Dengue is common in the tropics, but the risk varies from place to place depending on climate and other social and environmental factors.

According to one modeling estimate, 390 million dengue virus infections occur each year (95 percent credible interval 284–528 million), with 96 million (67–136 million) manifesting clinically.^1^ 3.9 billion people are at risk of dengue infection, according to another study. 129 countries have an infectious risk^2^.Asia bears 70% of the actual burden^3^.

In the previous 20 years, the number of dengue cases reported to WHO has gone from 505,430 in 2000 to over 2.4 million in 2010 and 5.2 million in 2019. Between 2000 and 2015, the number of deaths reported grew from 960 to 4032. Most of the deaths were in younger people. During 2020 and 2021, both the total number of cases and the number of deaths that were reported seemed to go down. But not all of the data is in yet, and the COVID-19 epidemic may have made it hard for some countries to record cases.

The dengue virus is transmitted primarily through mosquitoes. The mononuclear phagocyte cell lineage is where the dengue virus multiplies once it enters the body via the bite of an infected mosquito. Two uncommon forms of transmission are vertical and nosocomial. Blood transfusions, organ donations, and needle stick injuries can all spread Dengue, but this is highly uncommon. Dengue fever epidemics are becoming more frequent as the disease spreads rapidly to new locations. One in five blood donors are at danger of developing or being exposed to dengue fever, which has spread to more than half the world’s countries.

Before giving blood, blood donors are given a series of routine questions to help determine if they are in good health and clear of any diseases that could be transferred by blood transfusion. Donors are not permitted to donate blood if their answers indicate that they are ill or at danger of contracting a disease transmitted by blood transfusion.

Blood banks must have enough staff and a transfusion specialist on hand to ensure that the entire procedure is properly monitored. Because of a paucity of transfusion specialists in Bangladesh, this process is hindered. Furthermore, because of a lack of personnel, it is unable to conduct regular audits of the health authority. As a result, in our country, using questionairre’s criterion for donor selection has been fruitless. The primary goal of pre-transfusion screening is to avoid the transmission of infectious disease and to identify those who have already been affected. How might a patient and a donor profit from each other’s knowledge of their current health status. Dengue fever has become endemic in Bangladesh. Because most people with dengue are asymptomatic, it’s impossible to tell who’s already infected. A second infection might result in more catastrophic consequences such as dengue haemorrhagic fever and dengue shock syndrome.

This study aims to determine the percentage of donors who have dengue antibody in their blood when they donate it on a regular basis.

## Materials and methods

### Ethical approval

Before the study began, the Institutional Review Ethics Committee of Sheikh Hasina National Institute of Burn & Plastic Surgery gave its clearance. All blood samples were taken only after subjects had given their informed, written consent.

### Blood sampling and screening participants

This observational seroprevalence investigation was carried out at the blood transfusion service at the Sheikh Hasina National Institute of Burn & Plastic Surgery and the Dhaka Medical College Hospital. Between January and March 2022, 507 healthy Bangladeshi blood donors (aged 18 to 65) who were accepted for blood donation in accordance with the National Blood Transfusion Act’s guidelines and tested negative for the human immunodeficiency virus (HIV), hepatitis C virus (HCV), hepatitis B virus (HBV), syphilis, and malaria were randomly included. 3 mL of whole venous blood were drawn from each registered donor and put into BD vacutainer serum separation tubes. To collect serum, the tubes were centrifuged at 3,000 rpm for 5 minutes. Then, until they were screened, sera were kept in a freezer at −20 °C. Serum samples were tested using SD Bioline Dengue Duo for DENV non-structural protein 1 (NS1) antigen, anti-DENV IgM, and anti-DENV IgG antibodies at the conclusion of the collection phase. To identify dengue-specific antibodies, the SD Bioline Dengue DUO IgM/IgG test employs an immunochromatographic technique. Recombinant DENV-1, DENV-2, DENV-3 and DENV-4 antigens in the viral envelope bind to colloidal gold to create an antigenantibody complex. Human anti-IgG or anti-IgM immobilized in two discrete regions catch this complex as it migrates through the membrane of the test plate, resulting in recognizable pink bands in the appropriate places. Every sample tested negative for NS1 antigen.

### Statistical analysis

Data from the study’s sample were analyzed using SPSS 22.0. In total, 507 samples were gathered between January 2022 and March 2022 for the study. NS1 and IgM against DENV were both negative. So our main attention was on anti DENV IgG.

## Results

### Study participants

For the purpose of this research, a total of 507 people who donate blood were sought out. According to the data presented in Figure 1, a total of 93.3 percent (473 males) of the donors were male, while only 6.7 percent (34) were female. The overall population had a mean age of 27.7 years (standard deviation), and there was no significant difference between the mean ages of males and females (P = 0.506). The age groups ranging from 16 to 33 years and 18 to 25 years were the most represented among donors all across the world.

**Figure-1:**
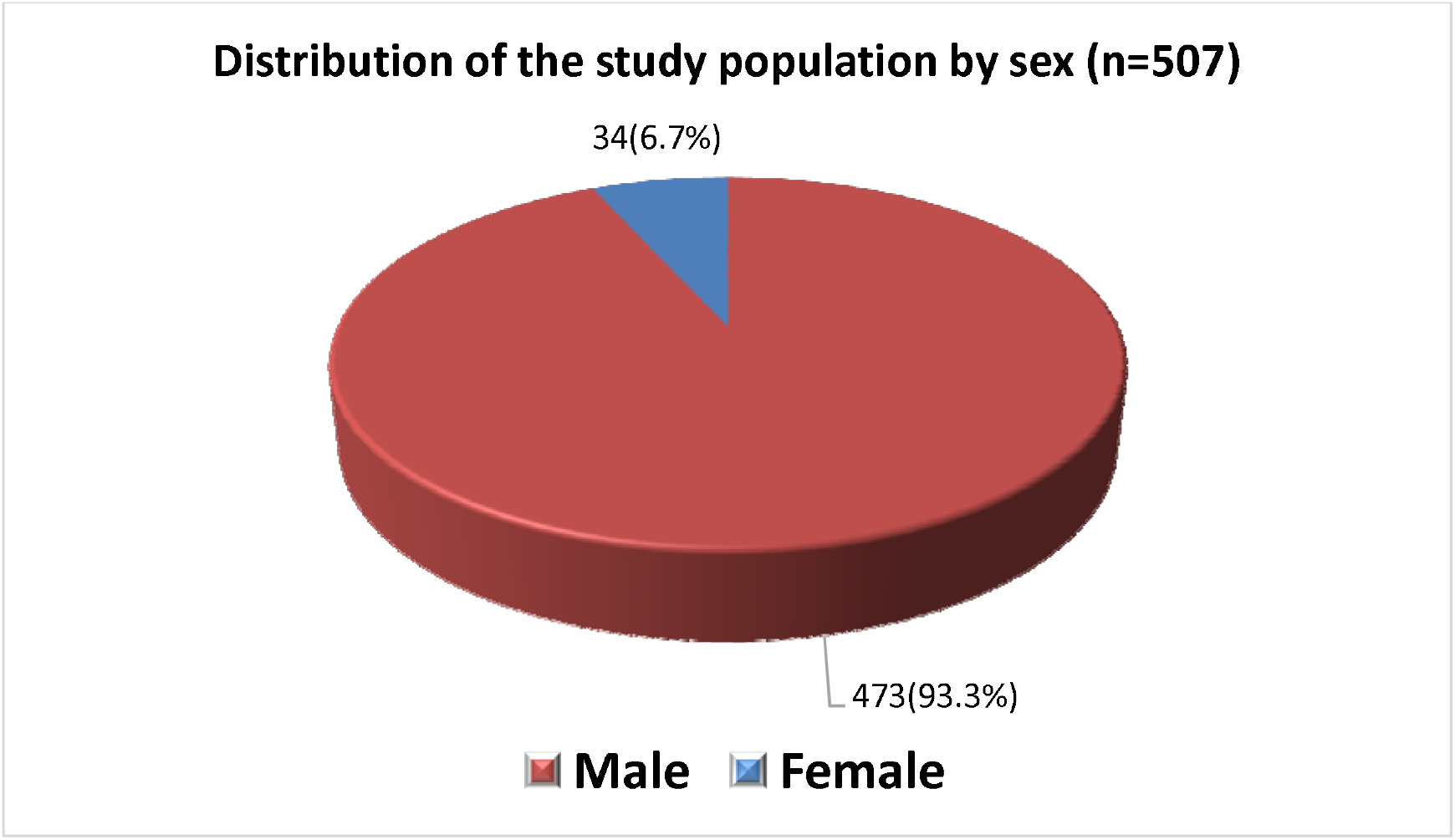
Pie diagram showing the sex distribution of the study population.

### Dengue virus infection overall prevalence in studied population groupings

According to the data presented in the Dengue Health Bulletin that was put out by the DGHS in Bangladesh, the months of June through September saw the highest number of probable DENV cases, with the peak occurring in the months of March and April. This coincides with the time of year when the study region experiences the most rainfall. Our research was carried out between the months of January and March, which is often considered to be the pre-monsoon season. During this time, the number of reported cases was significantly lower than in previous seasons.

A comparison between two groups using an unpaired student t-test was carried out in order to investigate the connection that exists between the various age groups and the test result. And the results showed that it was not significant.(Table-1).

**Table-1:** Association of age with dengue positive or negative cases (N=507)

| Age group<br>(years) | Dengue |  |  | p-value |
| --- | --- | --- | --- | --- |
|  | Positive<br>(n=183)<br>No. (%) | Negative<br>(n=324)<br>No. (%) | Total<br>(n=507)<br>No. (%) |  |
| <20 | 8(4.4%) | 9(2.8%) | 17(3.4%) |  |
| 20-29 | 114(62.3%) | 201(62.0%) | 315(62.1%) |  |
| 30-39 | 52(28.4%) | 92(28.4%) | 144(28.4%) |  |
| 40-49 | 8(4.4%) | 18(5.6%) | 26(5.1%) |  |
| 50-60 | 1(0.5%) | 4(1.2%) | 5(1.0%) |  |
| Total | 183(100.0%) | 324(100.0%) | 507(100.0%) |  |
| Mean±SD | 27.4±6.2 | 27.8±6.7 | 27.7±6.5 | 0.506<sup>ns</sup> |
| Range (min-max) | 18.0 – 50.0 | 18.0 – 60.0 | (18.0 – 60.0) |  |

Data were expressed as frequency and percentage and mean±SD
**Unpaired student t-test** was performed to compare between two groups
ns = not significant

The Chi-Squared Test (χ^**2**^) was carried out in order to investigate the connection between the test result and the gender difference(table-2). In addition to that, it found nothing important.

**Table-2:** Association of sex with dengue positive or negative cases (N=507)

| Sex | Dengue |  |  | p-value |
| --- | --- | --- | --- | --- |
|  | Positive<br>(n=183)<br>No. (%) | Negative<br>(n=324)<br>No. (%) | Total<br>(n=507)<br>No. (%) |  |
| Male | 173(94.5%) | 300(92.6%) | 473(93.3%) | 0.401<sup>ns</sup> |
| Female | 10(5.5%) | 24(7.4%) | 34(6.7%) |  |
| Total | 183(100.0%) | 324(100.0%) | 507(100.0%) |  |
| Male: female ratio | 17.3:1 | 12.5:1 | 13.9:1 |  |

Figures in the parentheses indicate corresponding **percentage**;
**Chi-squared Test ( $\chi^2$ )** was done to analyze the data. ns = not significant

Because the vast majority of blood donors are in their 20s and 30s, tests tend to focus on those donors’ blood.(Figure-2).

**Figure-2:**
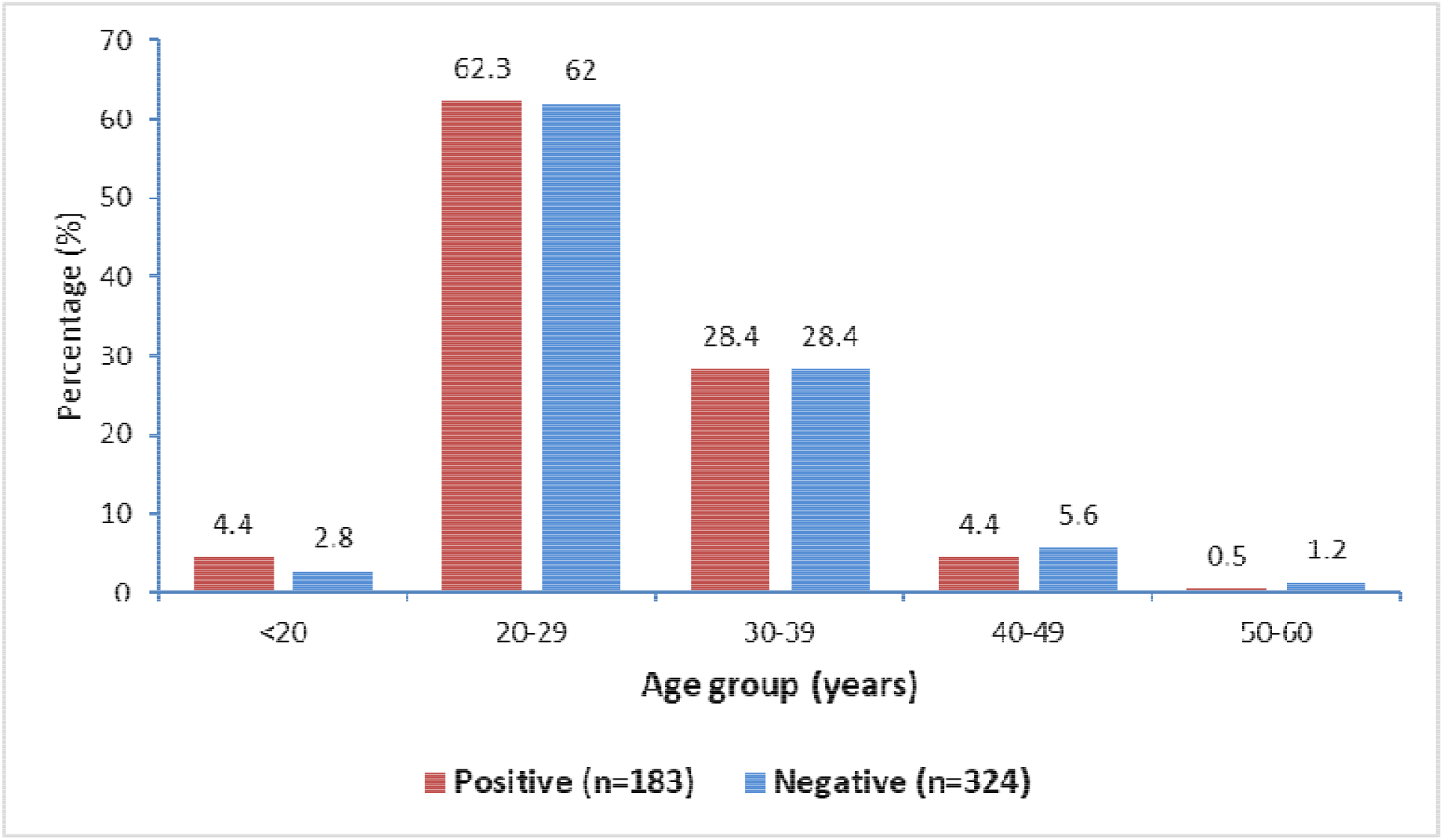
Bar diagram showing the age distribution of the study patients.

183 of the 507 samples produced anti DENV IgG positivity, while 324 did not. The ratio of positive to negative was 1.25:2(Figure-3).

**Figure-3:**
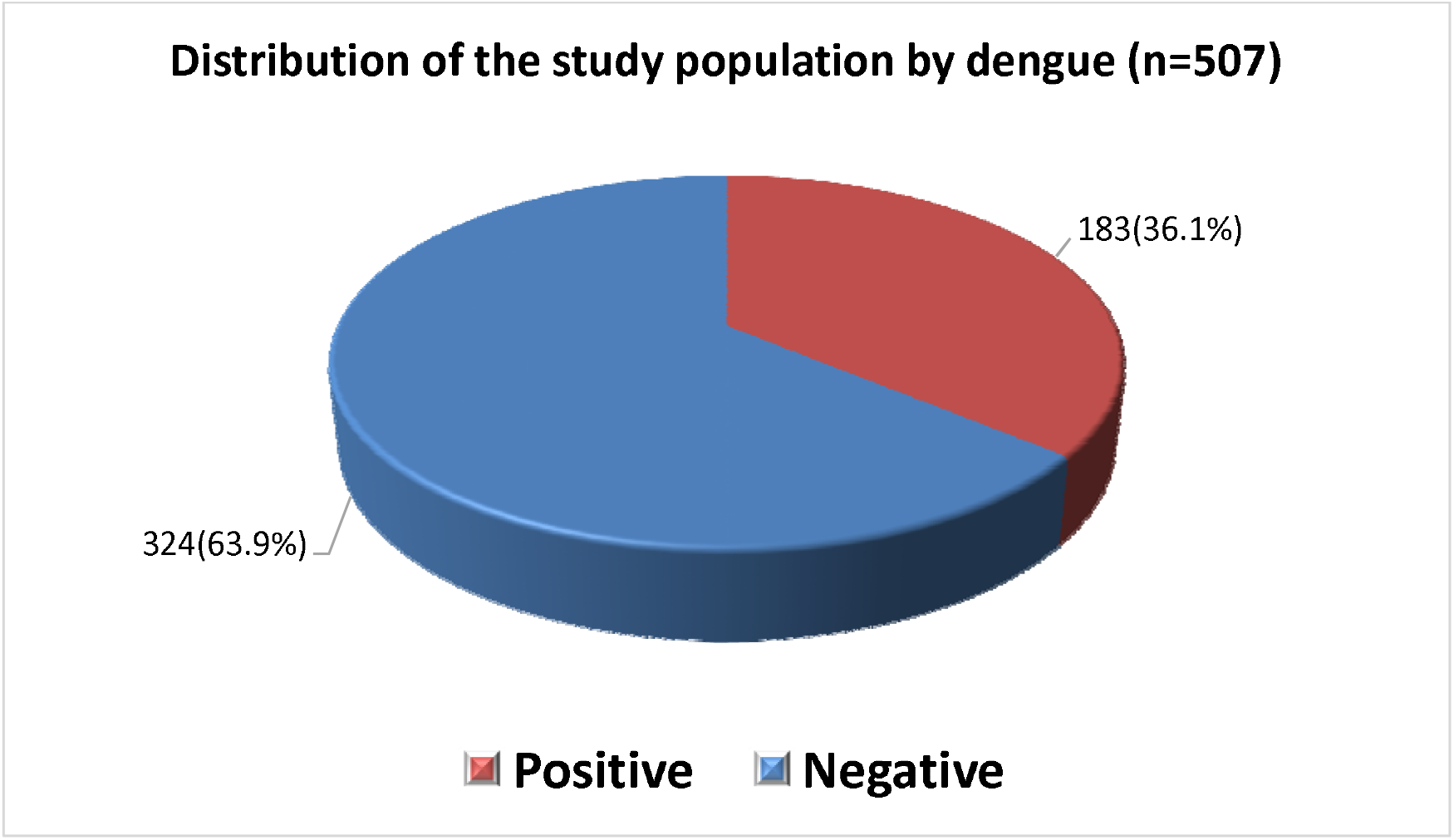
Pie diagram showing the dengue status of the study population.

## Discussion

Allogenic blood has never been more in demand as a resource than it is today ^4^. Escalating elective surgery, supply shortages, a lack of national blood transfusion services, policies, appropriate infrastructure, trained personnel, and financial resources to support the operation of a voluntary non-remunerated donor transfusion service, and old and emerging threats of transfusion-transmitted infection have all conspired to ensure that allogenic blood remains a vital but limited asset to healthcare delivery, particularly in the developing world.

Developing countries have significant challenges in providing a safe blood supply and blood transfusions. Because underdeveloped countries typically have insufficient available blood supply, they rely on family blood donors^5^. According to the findings of our research, around 36 percent of the donors who came to our centers to provide blood had positive anti-DENV IgG antibodies.

The same kind of research was done in Cameron and Central-West Brazil. Their respective results were 1.9 percent and 4.21 percent^5,6^. Four separate but related serotypes of the Flaviviridae virus cause dengue^7^. Infection recovery provides lifelong immunity to that serotype. Cross-immunity following recovery is partial and transitory. Secondary infections increase the risk of severe dengue^8^.

Defending against the dengue virus is mostly a function of the human body’s immune system. An infected person’s innate and adaptive immune responses work together to resist the dengue virus^9^. To combat dengue virus infection, the immune system’s B cells and cytotoxic T cells create antibodies that specifically target and destroy virus-infected cells. This phenomenon is known as “antibody-dependent enhancement,” and it occurs when a person gets infected with a different strain or kind of dengue virus and the body’s immune response makes the symptoms worse and raises the likelihood of getting severe dengue.

An antibody-dependent enhancement is another way that dengue virus infection can be made worse by antibodies. The dengue virus complex contains antigenically related viruses, yet more than one virus can infect a single host at a time. As a result, the secondary infection triggers a different antibody response than the main infection.

When the disease spreads across the population, the likelihood of receiving blood from a viraemic donor during the asymptomatic or subclinical phase of infection increases. The two reports from Hong Kong and Singapore, which both took place at the height of the outbreaks in these countries, show that dengue can be spread through blood from donors with no symptoms^10^.

Infectious and contagious agents can spread during blood transfusions because of delayed reactions that increase the risk of infection to recipients. Clinical and epidemiological screening of donors, as well as serological testing, ensures that blood is safe for transfusion purposes.

In spite of this, blood donors can still be exposed to infectious and contagious organisms through serological testing. The dengue virus is expected to be included to the serology of blood banks in tropical and subtropical countries in the near future. This is owing to the fact that 40% of dengue fever patients display no symptoms and so are not detected or reported to the health authorities^11^.

Non-neutralizing antiviral antibodies are present in people who have been infected with one of the four dengue serotypes. If the virus is re-infected, the antibodies will detect it but will not inhibit or neutralize it. Through opsonization, an antigen-antibody combination facilitates viral entry into macrophages and hence boosts viral replication. The macrophage releases vessel-active mediators, increasing vascular permeability and the risk of hypovolemic shock due to plasma leakage through the vessel walls.

To be clear, the risk of hemorrhagic dengue in those who get IgG anti-dengue antibodies is increased if they are infected with a second dengue serotype within six months of receiving the blood transfusion^12^. Heterophile antibodies from an earlier infection may make it easier for other virus serotypes to invade.

### Research limitations

1. A small number of people were included in the study.
2. No employment history was provided.
3. The PCR test was not performed.
4. The recipient’s follow-up was not carried out.

## Recommendations and Conclusion

1. A research should be conducted to assess the actual situation in a country like Bangladesh that adheres to WHO guidelines for transfusion screening but does not regularly administer anti-DENV.
2. It is possible to perform routine tests of anti-DENV IgM and IgG antibodies if the prevalence is high. A positive donor could result in a delay for a set period of time, such as six months, which the local government could put into effect.

## Data Availability

All data produced in the present work are contained in the manuscript

